# A scoping systematic review of implementation frameworks to effectively transition interventions into clinical practice in oncology, nuclear medicine and radiology

**DOI:** 10.1101/2022.03.05.22271946

**Authors:** Gayathri Delanerolle, Heitor Cavalini, Peter Phiri, Leslie Gelling

## Abstract

**Background:** Clinical research studies have made significant strides globally requiring clear processes to transition research interventions into clinical practice. Theoretically, implementation of novel interventions require clear methods as part of a fit-for-purpose (FFP) framework comprising of effective adaptation processes in conjunction with practice policies. Implementation science (IS) based operational research (OR) is vital in global health as it addresses the ‘know-how-do’ gap using a real-world setting to achieve best practices to sustain healthcare. Despite this, limited OR is available to evaluate and validate implementation frameworks for complex clinical specialties such as oncology, diagnostic radiology (DR), nuclear medicine (NM) and interventional radiology (IR). This is the first study to systematically review implementation frameworks including its’ validity and applicability in healthcare.

**Method:** We searched 17 databases including PubMed, Medline/OvidSP, Science Direct, PROSPERO, PRISMA, PubMed Health, Embase, EBSCOhost, SciELO, TRIP, ProQuest, Academic search complete, Ageline, Cochrane, Web-of-Science and BIOSIS using a comprehensive search strategy and MeSH indexing to review publications from January 1^st^ 1980 to 31^st^ March 2019 in English. We selected 20 publications as per the inclusion/exclusion criteria developed under a review protocol registered with PROSPERO (CRDG42019124020).

**Findings:** There were no publications indicating a validated framework or a specific system used to implement evidence based interventions (EBIs) within oncology, IR, NM and DR although there were generalized implementation processes, adaptation models and policies. Furthermore, validation studies were not conducted against these frameworks to review their applicability and viability in healthcare especially in the UK.

**Interpretation:** It is evident there is a research implementation gap in healthcare and further research is required to establish a fit for purpose framework to cover multiple ‘blind spots’ using a real-world (RW) setting. Current evidence also suggests, alignment of academic theories to healthcare including its applicability to various clinical specialties is needed.

## Introduction

Clinical research is considered as a cross-disciplinary specialty in modern day medicine that merges science and clinical arenas to develop treatments, methodologies and medical technologies to better public health (CRUK 2018; Garralda *et al*. 2019). The rapidly changing clinical research landscape has led to an array of clinical treatments that are moving towards a precision medicine platform (Choi *et al*. 2018, Garralda *et al*. 2019) giving rise to complex interventions (CI), especially in oncology, nuclear medicine (NM), diagnostic radiology (DR) and interventional radiology (IR). CIs vary across clinical specialties thus, a detailed summary of those relevant to this study has been listed in table 1.

**Table 1.**
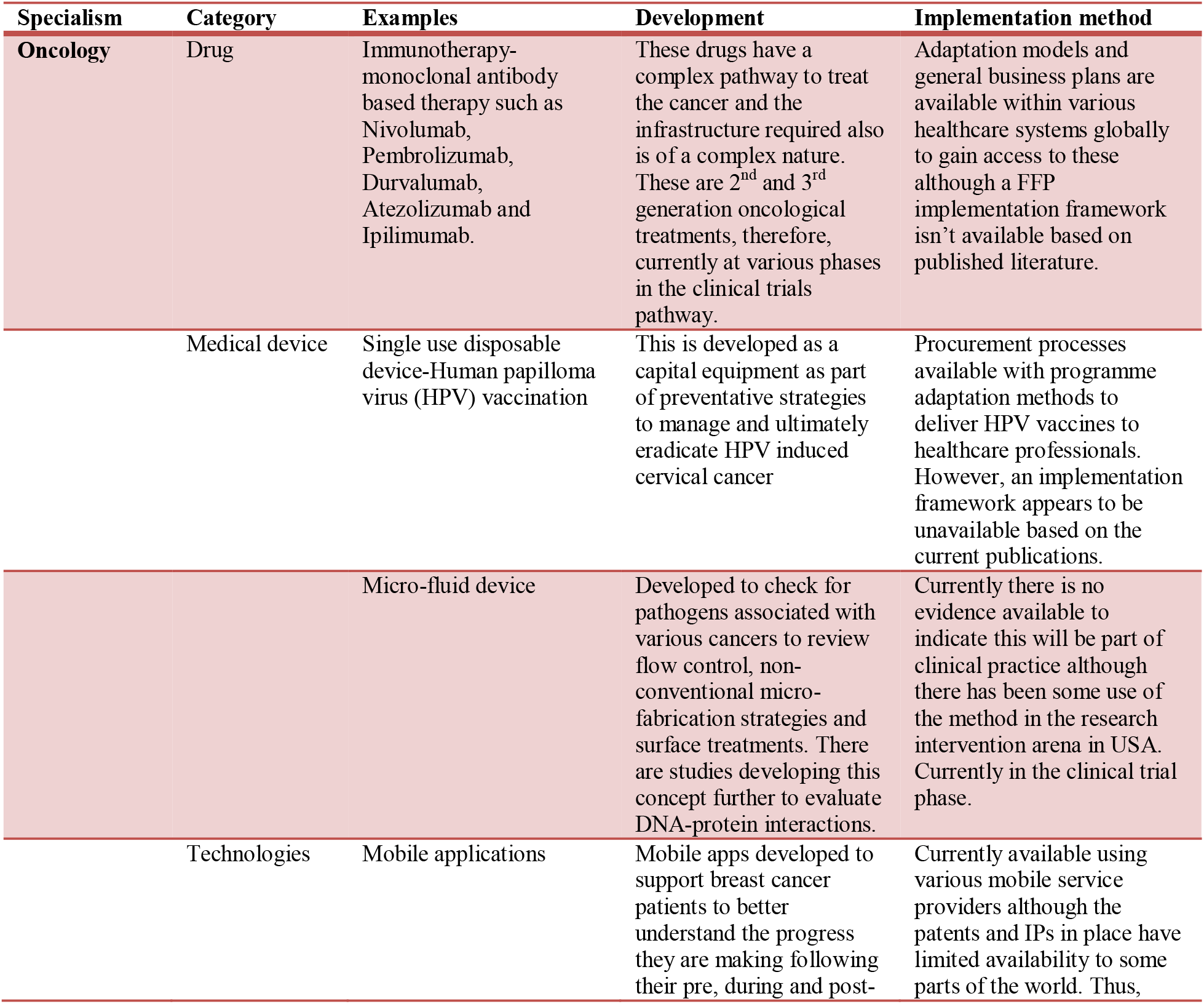
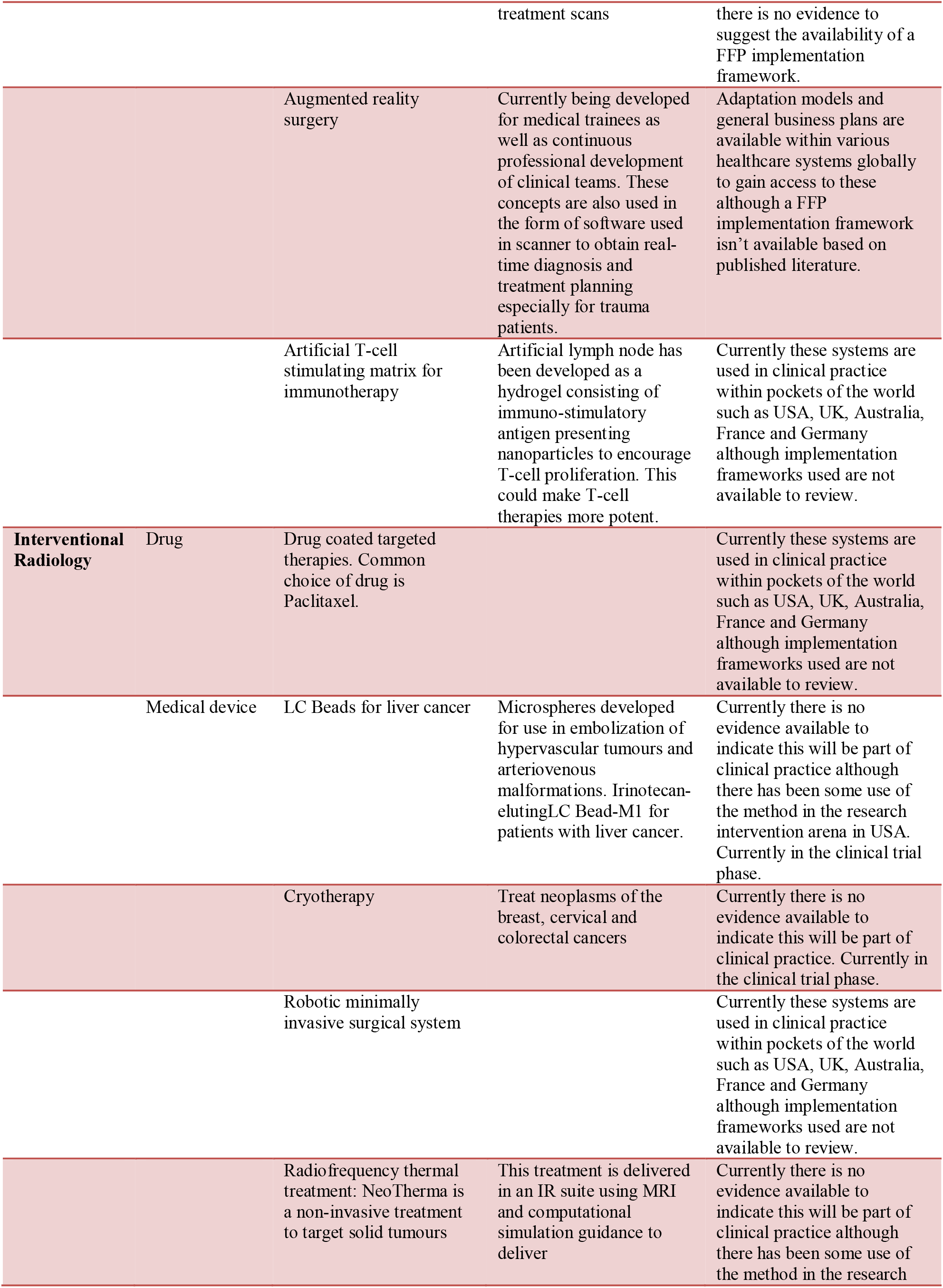
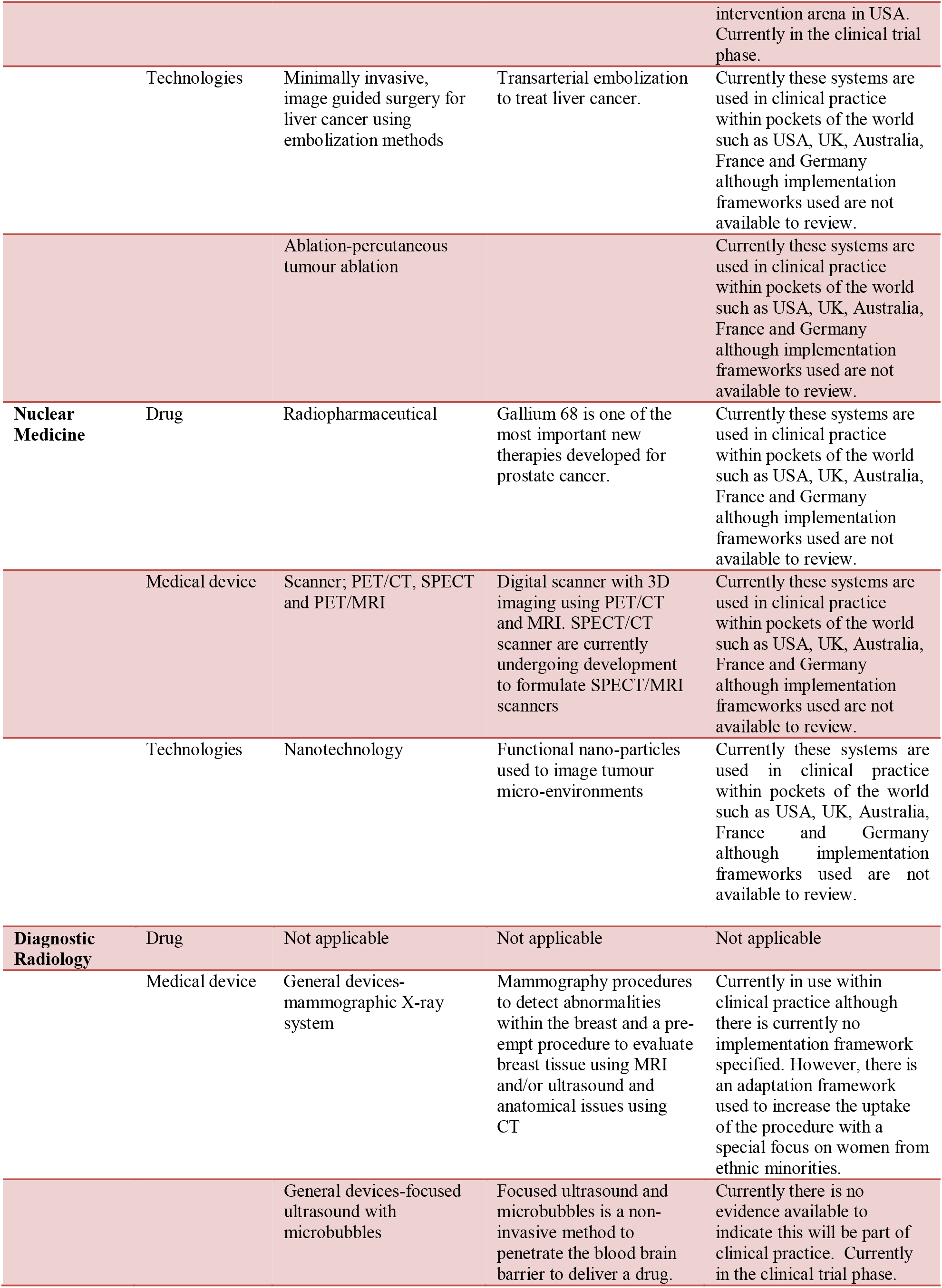
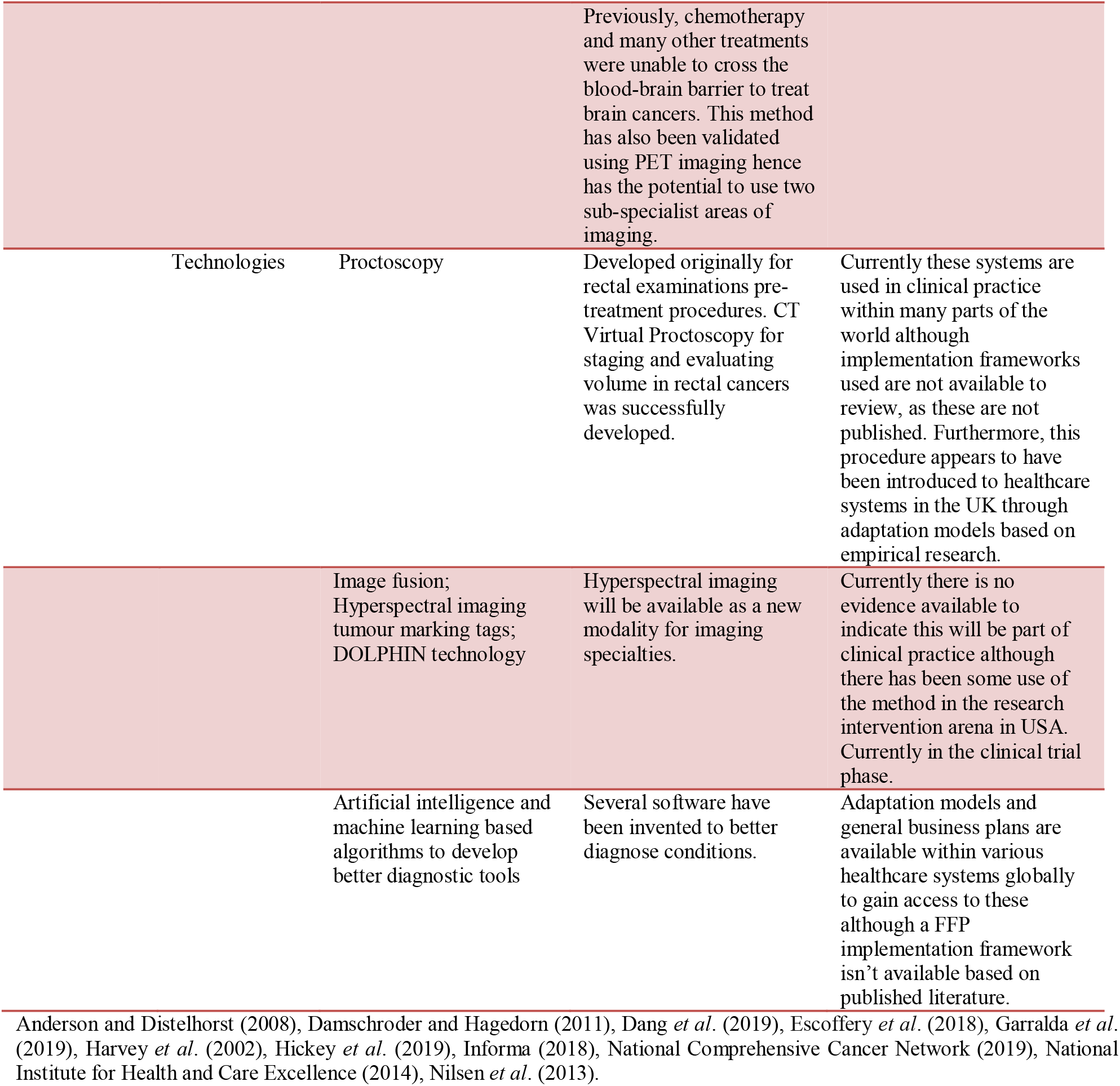
Complex interventions (CI) within clinical specialties

According to the world health organisation (WHO), the cancer incidence rates will continue to increase globally whilst the cancer registry in the UK reported 303,135 new cancer cases in 2016 (CRUK, 2018). The WHO attributed 9.6 million global deaths to cancer in 2018. Therefore, the importance of delivering high quality oncological care is vital to maintain the ‘covalent bond’ shared between research innovation and the clinical interphase (Garralda *et al*. 2019, Harris *et al*. 2009). Within the last decade, oncological care has become heavily dependent on NM, DR and IR for diagnostic and treatment purposes. Despite these significant strides in precision medicine, there appears to be challenges in translating research outcomes to policy and clinical practice (CRUK, 2018, Franks & Schroeder 2013, Garralda *et al*. 2019). This also includes challenges within workforce training practices (Chambers & Norton 2016, McKleroy *et al*. 2006, Montez *et al*. 2016) which indicate a significant variability in knowledge management and transfer within healthcare as indicated in table 2.

**Table 2.**
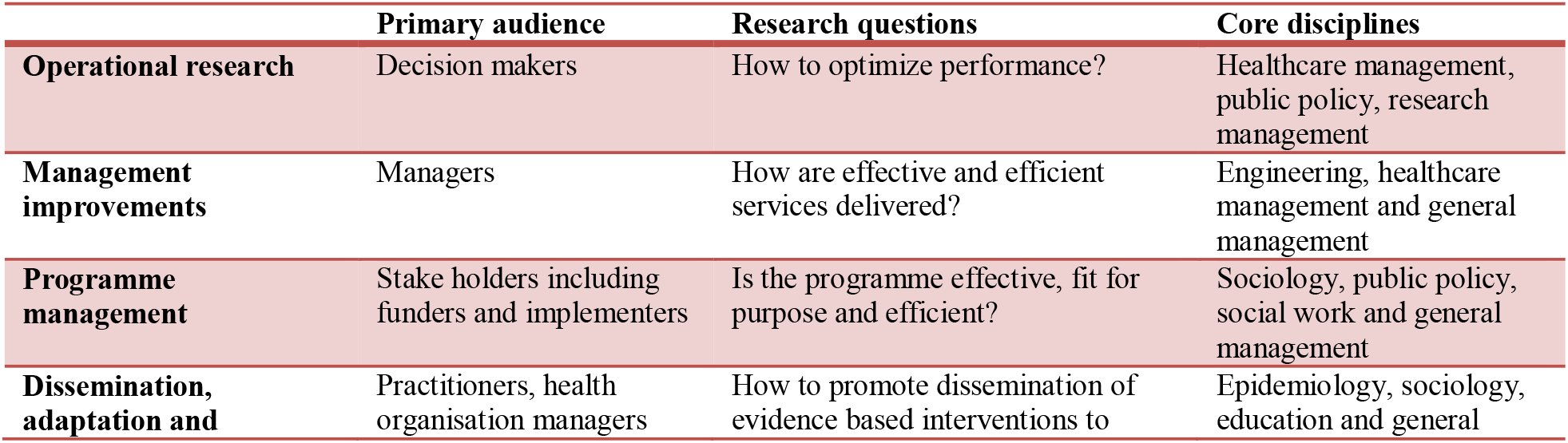
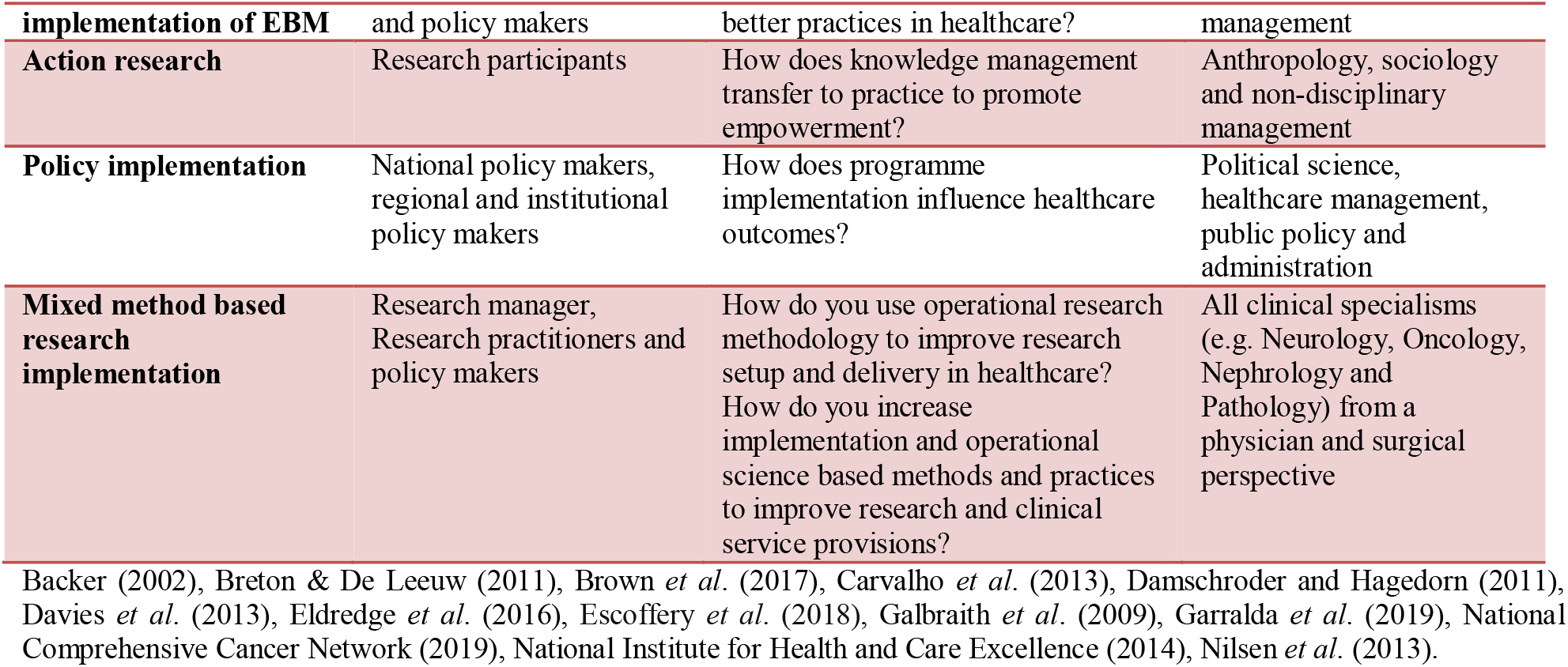
Implementation research facets with targeted audience

Implementation science (IS) has the ability to address these challenges using an evidence-based practice (EBP) approach (Durlak 2015, Escoffery *et al*. 2019, Mailk *et al*. 2018). Some implementation practices show success through empirical driven concepts instead of published theories (Madon *et al*. 2007). Eccles and colleagues (2005) indicated that this type of research appear to be ‘an expensive version of a trial-and-error method’ (Eccles *et al*. 2005) whilst Davies and colleagues (2003) stipulated that only 10% of studies identified guidelines for implementation strategies with an explicit rationale but provided no evidence to indicate these were effectively used in any healthcare setting (Davies 2013). However, there is a general acceptance that implementing EBP in different environments attributed to limitations in the theories used (Nilsen 2015). In addition to this, there appears to be poor theoretical underpinning to track the failure of implementation strategies and their precise reasoning, thus, preventing opportunities to better predict and formulate a series of processes as part of a framework that is verified and fit-for-purpose (FFP).

Analysis of the global drug (Medicines & Healthcare products Regulatory Agency 2014) and device development landscape indicates that oncology has the highest therapeutic class in terms of clinical innovations and specialization, thereby, directly influencing innovations and practices in NM, IR and DR due to its/ intra-dependency (Clinical trials NHS 2018, European Medicines Agency 2019). Hence, it isn’t a surprise that oncology as well as imaging research is considered highly complex (Malik *et al*. 2018, Medicines & Healthcare products Regulatory Agency 2014). Study complexity varies depending upon the type of drugs or interventions developed, regulatory requirements, treatment demand, development time, research and developmental costs, patient expiration, globalization and the implementation into clinical practice (International agency for research on cancer 2017, Medicines & Healthcare products Regulatory Agency 2014). Furthermore, fast-track approvals of new therapies (EAMS, 2019) have also been discussed in detail in the US (Moore *et al*. 2013, National Cancer Institute 2018) and Europe (EAMS, 2019), thereby supporting the need for FFP implementation procedures to succeed ensuring timely access for patients. Therefore, the responsibility of conducting research and its’ adaption into healthcare has become a vigorous exercise on a global scale (Bartholomew *et al*. 2001, Castro *et al*. 2004).

In addition, maintaining pace with the changing research landscape with rising CIs and increase in treatment demand due to an aging population and increases in incidence of cancers (Clinical trials NHS 2018, CRUK 2018, International agency for research on cancer 2017), it has become challenging to promote logarithmic growth in the UK as the current NHS infrastructure is consistent with rate limiting factors involving infrastructure (CRUK 2018). In a bid to support the much needed infrastructure, healthcare organisations consist of a clinical and research service provision especially in the UK with shared resources (Chambers & Norton 2016, CRUK 2018, Escoffery *et al*. 2019). Furthermore, the National Institute of Health Research (NIHR) was established as the ‘research arm’ of the NHS, centrally coordinating research setup and delivery between industry, academia and NHS organisations. In addition to the NIHR, the Health Research Authourity (HRA) supports research study setup by coordinating between industry, academia, the NHS and national ethics committees using a multitude of processes (Clinical trials NHS 2018, Cooke 2005). The responsibility of delivering research care is entirely at an organisation level (Clinical trials NHS 2018) and it is evidence based on the NIHR recruitment matrix, all NHS organisations show a commitment to conducting research which is also supported by the NHS constitution.

Although, the NIHR report increases in research activities, clinicians, researchers and policy makers continue to equally report of challenges to conduct research and translate outcomes to clinical practice in a timely manner (Moore *et al*. 2013, National Cancer Institute 2018). These barriers include time taken to conduct clinical trials, complexities in procedures, funding concerns, delays in publishing results and influencing governing bodies as well as organizational cultural issues (Dilts *et al*. 2009, Franks & Schroeder 2013). Dilts and colleagues (2009) reported process complexities that indicate approximately 300 distinct processes associated with activating a phase III study and an associated median time from conception to activation being approximately 600 days (Dilts *et al*. 2009). Furthermore, trial outcome reporting was explored by Ross and colleagues (2012) indicating the overall publication rate to be 68% whilst Goldacre and colleagues (2018) reported this to be 49.5%, highlighting majority of these are from commercial sponsors. Although the difference in the rates of publications could be attributed to changes in the number of studies explored and difference in time frames, it still indicates research performance by way of reporting outcomes has reduced despite increases in the number of clinical trials being conducted globally (Choi *et al*. 2018). Based on this collective evidence and in comparison to high activity reporting from the NIHR, it is clear that current processes require improvements for effective implementation of interventions into clinical practice. Furthermore, it also evident, better strategies are required to promote transparency into research outcome reporting leading to the construction of better adaptation models as part of an effective framework especially for Cis (McKleroy *et al*. 2006, Moore *et al*. 2013). Also, to promote evidence based practices/interventions (EBP/EBIs), dissemination and implementation processes need to be more synergistic across research feasibility, setup and delivery (Castro *et al*. 2004, Franks & Schroeder 2013, McKleroy *et al*. 2006). There, is also an added complexity in that currently, there is limited research in IS especially in the UK compared to the US for example (International agency for research on cancer 2017, Krivitsky *et al*. 2012). Therefore, improvement and implementation science based research within complex clinical arenas such as oncology and imaging is nascent. It is also unclear thus far, if general framework theories identified and published is useful and effective within complex specialties as there is lack of evidence to indicate the validity and viability within healthcare systems such as the NHS organisation. In order to address this issue, extensive operational research based studies are required. Thus, the purpose of this systematic review is to use scoping methodologies to identify and summaries the systems and/or frameworks currently available to promote EBP in oncology, IR, NM and DR. The study also aims to review any empirical research and grey literature from an operational performance outcomes perspective and any published cultural paradigms in implementing EBIs in a healthcare setting.

### Panel 1: Research in context

#### Evidence before this study

We performed a thorough search of the scientific literature using Pubmed, PROSPERO, Medline and Cochrane Reviews prior to undertaking this study. The initial review indicated there are challenges around implementation of research interventions within clinical practice due to a variety of challenges. It also indicated implementation science is at its foetal stages within many clinical specialties in the UK. Challenges were reported via empirical and grey literature. There was minimal implementation research available to use within the context of the UK healthcare system that focused on IR, NM, DR and oncology, indicating a potential knowledge gap and a conjugate-bridge between theory, its’ practice and validation. With growing healthcare demands, a robust implementation framework is required to deliver quality clinical care safely and minimize research waste. Therefore for the purpose of this study, we have considered the following implementation characteristics to be used as part of the framework;

- ***Context specific***
- ***Fit for purpose and demand driven***
- ***Real world and real time***
- ***Focuses on processes and outcomes***

#### Implications of all the available evidence

The current evidence postulated from this study indicates the need for further research to obtain an implementation framework that would be purposeful to oncology, IR, NM and DR enabling a smoother transition of complex interventions from the research arena to clinical practice. Furthermore, it is also evident, better practice policies to mitigate barriers to implement and uptake of novel interventions is required. In addition to this, further knowledge is required to bridge the *‘theory and practice*’ gap within implementation science, as currently, most published literature appears to lack validation methods.

## Method

### Added value of this study

This study aims to review the current published literature that indicates implementation frameworks globally available within healthcare and/or clinical specialties. This is the first systematic review conducted to evaluate implementation frameworks used in CIs. Outcomes from this study could optimise operational performance and establish relevant processes and policies to improve inter and intra-disciplinary workings in oncology, IR, NM and DR practices to better deliver and use clinical trials to enhance clinical practices.

### Search strategy and selection criteria

The methodology was developed as part of a protocol that is consistent of a structure approved by the NHS centre for reviews and disseminations managed by University of York and the NIHR. This systematic review was registered with the international register for systematic reviews PROSPERO: CRDG42019124020. The developed protocol provides transparency, reproducibility and transferability to promote good practices.

The systematic strategy developed used literature search engines such as Medline/OvidSP, Science Direct, PROSPERO, PubMed, PubMed Health, Embase, EBSCOhost, SciELO, ProQuest, Academic search complete, Ageline, Google Scholar, PRISMA (preferred reporting items for systematic reviews and meta-analyses, Cochrane, Web-of-Science and BIOSIS. These databases were searched until the 10^th^ of June 2021. The search was conducted using a ‘shot gun’ explosions and combinations approach along with the medical subject headings (MeSH) indexing method. Using the MeSH method, a multi-text classification approach was used with key phrases and words such as adaptation research, implementation science and research, in oncology, implementation research in imaging, translational framework for research delivery in oncology, research systems, evidence based policy, evidence based health services research, research culture impact in implementation and research delivery framework. Within the context of this study, specific definitions were also developed and used for key terms of “*frameworks, model, research capacity-capability and theory*”, which have been described in Appendix 2.

As part of the systematic methodology, an inclusion and exclusion criteria was developed to generate a bibliography of candidate studies. At the onset, the process of selection was evaluated by piloting the inclusion/exclusion criteria on a subset of studies as indicated in figure 1. A second independent evaluator reviewed the reliability of the initial finding. The inclusion criteria included papers, reports and grey literature in English, published in journals and reputable websites which indicated research processes and/or systems associated with delivering research outcomes in a healthcare setting, specifically in oncology, IR, DR and NM. Furthermore, publications which indicated an implementation framework based on empirical research, implementation models and any other compliance driven frameworks or policies were also included if there was evidence to indicate their validation by any healthcare organisation. Publications in the form of commentaries and professional opinions were included with the exception of those without research and/or clinical evidence to support any conclusions.

**Figure 1.**
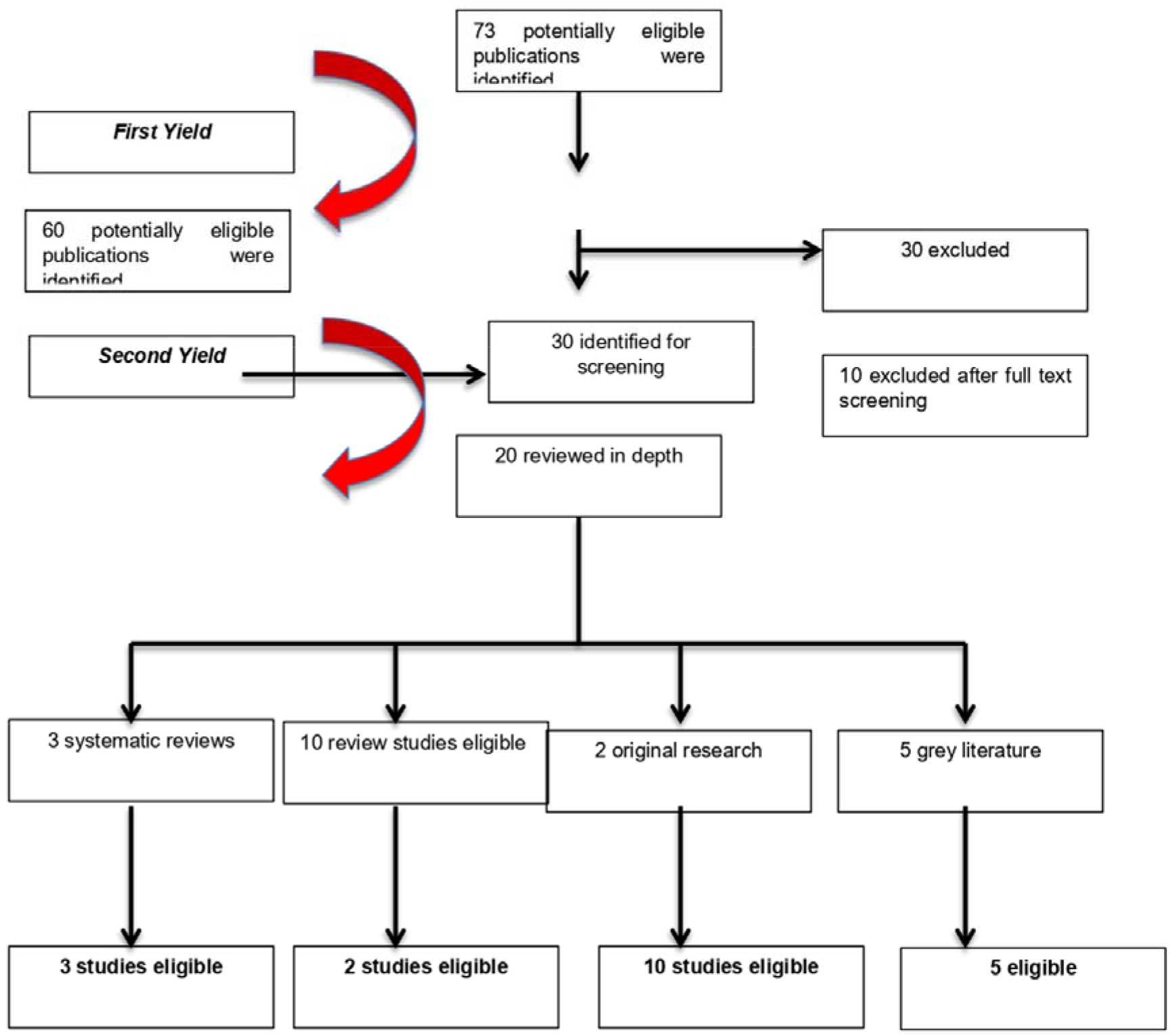
Shows the study review procedure used

### Quality assessment of reviews

One reviewer and checked/verified extracted data by another from publications, which was crosschecked for accuracy. Three main types of data were extracted which includes methods, characteristics of operational frameworks and improvements. Quality assessment of the material reviewed is an important aspect to consider. Therefore, the strength of evidence presented in each of publication was considered on an individual basis. The quality as well as the scope of the publications used varies widely. AMSTAR was used to assess methodological quality of this systematic review (Ayres *et al*. 2005).

## Results

Locating the most appropriate literature for this subject matter was challenging. As this is a primary research review, the paradigm used was a mixed methods approach. Thus, exploring the current level of general practice knowledge available and published in other clinical areas such as immunology and mental health. The results were based upon on reviewing the data with 3 levels of knowledge synthesis of system, correlation and descriptive analysis. Of the 30 studies, there were no studies published that were directly relevant to oncology, IR, DR and NM. However, 20 publications with adjunct text were found that indirectly showed key characteristics required to cultivate adoption of EBIs within a clinical service provision. As there was limited data available, a meta-analysis was not conducted to synthesize the research findings. A narrative synthesis with specific outcome measures as per the protocol designed was used to review the 20 publications that met the inclusion criteria.

We identified 20 frameworks although only 2 have a direct association with the research question. Out of the 20 frameworks identified, 13 included adaptation steps from gray literature specific to the US (Backer 2002, Bartholomew *et al*. 2001, Chen *et al*. 2013, Eldredge *et al*. 2016) and 7 from published literature (Bartholomew *et al*. 1998, Chen *et al*. 2013, Eldredge *et al*. 2016). Although, some aspects of these publications could be transferrable, it is evident the conclusions are dialectic with mutual citations as well as acknowledgment of building new frameworks based on previous work using adjustments thereby diluting any meaningful applicability to complex specialist areas such as oncology, IR, DR and NM. Furthermore, vast majority of this interconnectedness is applicable in the US as the frameworks developed were supported mainly by US government agencies such as the National Institute of Allergy and Infection, National Institute for Child Health and Human Development, Centre for Substance Abuse Prevention and Centers for Disease Control and Prevention (Krivitsky *et al*. 2012). This further justifies the frameworks developed have used the specialties of HIV (Galbraith *et al*. 2009), infection control and pregnancy and any direct applicability to other specialties would be negligible. The use of intervention mapping (IM) to adapt EBIs is becoming more common as indicated through empirical research and case studies reviewed. Although, IM indicates to be a useful tool for increasing uptake of a particular intervention and evaluating EBIs such as mammography, it remains to be seen if this method remains purposeful for CIs, as currently there is a lack of published data (Harris *et al*. 2009).

## Discussion

The systematic evaluation stipulates the need for a more formative and constructive implementation framework for CIs within oncology, IR, DR and NM. It is evident, there is a lack of data available to enable meaningful use of frameworks available. Current theories and concepts that could provide a meaningful framework and subsequent policy to implement EBIs rapidly is rate limiting. Currently, implementation of novel programs appear to use practice facilitation and learning collaborative initiatives to promote implementation strategies using organisation structures and workflows despite being labour intensive to staff and minimal service improvement as per patient reported outcomes (Ayres *et al*. 2005, Harvey *et al*. 2002). The costs of using these 2 methods also hinder widespread adoption and inclusion into implementation strategies, frameworks and any logic models used within research management structures. However, implementation tools that are cost effective and electronically available with tailour made options provide better efficiency and consistency to adopt site-specific requirements thereby increasing the uptake of the strategy. This structured approach could be proactively shared between a clinical and research service provision thereby sharing data more readily. However, regulatory restrictions within research may require additional steps to be included into the electronic system to adhere to research governance policies (Johnson *et al*. 2007, Medicines & Healthcare products Regulatory Agency, 2014).

Cole and colleagues (2015) indicate the use of an implementation strategies based adaptation programs for their mail based colorectal cancer screening service which identifies a consolidated framework for implementation research (CFIR). This framework highlights 5 factors within the model; intervention and individual characteristics, inner and outer setting and processes, which influence success of implementation when translating EBIs (Choi *et al*. 2018). The CFIR method has also been used to guide adaptation and evaluate EBP programs for substance abuse (Damschroder & Hagedorn 2011, Hsiao *et al*. 2013). However, generalizability of the CFIR in the context of complex interventions may have rate limiting factors within a secondary care setting in the UK given Cole and colleagues (2015) conducted their research only in 3 out of 7 centers in a primary care setting without any statistical significance (Choi *et al*. 2018).

Another facet to IS is dissemination and implementation (DAI) (Nilsen 2015). DAI is composed of single concept models than a framework. DAI is a fairly new area of operational research in which EBI adaptation is observed using multiple processes in various settings (Nilsen 2015). This concept has been used to build multiple taxonomies to adapt programs to implement EBI using modifications including lengthening, shortening, removing, substituting, addition, re-ordering program components, integration of multiple EBIs, loosening structures or departing from the EBI completely as discussed by Stirman and colleagues. On the contrary, Chambers and Norton indicated this ‘adaptome model’ enables knowledge synthesis in regards to program adaptation and its impact on implementation as well as outcomes thereby could function without a framework (Chambers & Norton 2016). There are however, debates in relation to fidelity versus adaptation whilst there is also evidence to suggest some level of adaptation occurring via empirical research, which isn’t often published (Carvalho *et al*. 2013). Furthermore, this data indicates these require validation in the context of CIs. The tension between adaptation and fidelity resulted in adaptation frameworks stipulating steps institutions should take to select and implement EBI to make the program (Bartholomew *et al*. 2001, Carvalho *et al*. 2013, Harvey *et al*. 2002). The converse could also occur if the most FFP context is not used, any changes made could make the EBI ineffective (Hsiao *et al*. 2013, Montez *et al*. 2016). However, common denominators of multiple frameworks with systematic processes could influence organisations to adapt any program. By way of these theories, frameworks and models, D&I research could improve EBI delivery and maintain long-term sustainability and increase rate of acceptability (McKleroy *et al*. 2006). Wingood *et al* (2008) and McKleroy *et al* (2006) indicates researchers and practitioners recommend frameworks to guide implementation of Cis (McKleroy *et al*. 2006, Wingood & DiClemente 2008). There are a limited number of D&I models and frameworks published and used in healthcare globally but the absence of a comprehensive review of processes and systems used in adaptation processes and practices within the literature prompted Bartholomew and colleagues original study (Bartholomew *et al*.1998).

Furthermore, processes and their systems associated with delivering research and its outcomes as well as its’ transition into clinical practice differ across institutions and continents. Publications from the US show organisations are likely to be conducting D&I on an ad-hoc basis which is driven by the requirements of the population size and oragnisational capacity (Carvalho *et al*. 2013, Castro *et al*. 2004, Galbraith *et al*. 2009, Hsiao *et al*. 2013). This further purports, a need for a specific framework to be introduced for CIs seen within cumbersome clinical areas. Although dissemination of research is vital to ensure EBI could be implemented to improve clinical practice, the adaption process can be complex and laborious given the variables observed across clinical specialties as well as healthcare organisations and their country of origin. Some researchers state adaptations is a vital and natural ‘next step’ as part of the implementation process to clinical practices, which could be further influenced by modifying the original CI, although this could reduce the efficacy and effectiveness (Bartholomew *et al*. 2001, Brown *et al*. 2017, Johnson *et al*. 2007, Krivitsky *et al*. 2012). Currently, most of the frameworks published for D&I are specific to the USA and specialties such as HIV, mental health, pregnancy and infection control specifically for diseases such as Malaria (Escoffery *et al*. 2018, McKleroy *et al*. 2006, Wingood & DiClemente 2008). Escoffery and colleagues conducted the first systematic research study, which showed a summary of adaptation frameworks for public interventions that can be used in practice highlighting adaptation tasks and steps as part of a framework (Escoffery *et al*. 2018). The study highlights frameworks to guide healthcare staff working in HIV, pregnancy and substance abuse prevention domains (Eldredge *et al*. 2016, Escoffery *et al*. 2018, Escoffery *et al*. 2019). However, the frameworks used were validated using the US healthcare system, which limits its use. Furthermore, the study lacked evidence to show it’s applicability in CI implementation.

Although ‘one size does not fit all’, validated FFP frameworks are required to better policy and practice within healthcare. This is further substantiated by several European governments acknowledging the implementation policy gap that is to improve healthcare practice (table 3). The mixed method systematic review approach used in this study indicates minimal research was available. Therefore, comprehensive research is required to better understand specialty based needs to design sustainable and FFP frameworks to enhance clinical practices.

**Table 3.** Key features published in policies associated with implementation science based research

| Evidence of research policy use in different areas |  |  |
| --- | --- | --- |
| Policy | Methodology preference | Evidence base |
| Healthcare (including NHS and World Health Organisation) | The current gold standard is systematic reviews although this is primarily appropriate for non-interventional research. This approach could be used retrospectively as well as prospectively. Systematic reviews could also include a multitude of sampling and synthesis methods using a single or multiple protocols (inclusive of longitudinal sampling). | 1. Systematic reviews can be expensive if conducted along with longitudinal studies via Cochrane collaborations and/or NHS clinical effectiveness unit data evaluation strategies. However, not all systematic reviews are expensive to conduct especially if it is conducted using applied or operational science models. |
|  |  | 2. Systematic reviews are also used in the context of 'real-world' data in conjunction with meta-analysis methods to aggregated statistics approaches. |
|  |  | 3. Systematic reviews provide evidence for low intensity support services, which include publications that have |
|  |  | methods poorly developed. However, this written evidence is strengthened with the inclusion of empirical knowledge and phenomenology. |
|  | The gold standard for novel therapeutic interventions are randomized controlled trials (RCT) inclusive of qualitative and socio-economical modeling methods to provide a comprehensive overview of the efficacy of the therapy explored as well as the views of the participants. This is often referred to as mixed-methods based RCT. This method could be used prospectively only. | 1. These studies are expensive and often require a great length of time to collate as well as analyse the data. They can often use existing data via Cochrane collaborations as part of the RCT analysis method to further the evidence base. |
| **Social care** | Current gold standard is to use qualitative methods, which includes quantification and experimentation with minimal evidence. Often qualitative research is analyzed in a narrative format, which is viewed with suspicion and even hostility (). | 1. Care data database of research is available via the Social Care Institute for excellence. This data is outside of the NIHR's clinical trial gateway database making it challenging to draw conclusions for clinicians or even use within existing diagnostic or screening based studies. |
|  | There is growing evidence to indicate social care methods are being used as part of mixed method approaches in the context of operational and applied science based research. The benefits of this approach enhances the manner in which interventional based clinical research is conducted and delivered in addition to the optimization of service | 2. Evidence using these methods is now being published in clinical and applied science journals, which is allowing a broader use. |

Breton and De Leeuw (2011), Clinical trials NHS (2018), CRUK (2018), Krivitsky *et al.* (2012), Mayer (2017), Montez *et al.* (2016), National Cancer Institute (2018).

## Data Availability

All data produced in the present study are available upon reasonable request to the authors

## Notes

### Competing Interest Statement

The authors have declared no competing interest.

### Funding Statement

This study did not receive any funding

